# Integrative Profiling of Glymphatic Dysfunction in Adolescent Subthreshold Depression

**DOI:** 10.64898/2025.12.18.25342437

**Authors:** Qiwei Guo, Ruisi Wang, Zhongli Lan, Zihan Wei, Utteng Sou, Chengfeng Zhang, Yueqi Zhang, Xingwei Fang, Jingli Zhang, Min Lu, Andriy Myachykov, Zhen Yuan, Jun Chen

## Abstract

**Objective:** Subthreshold depression (StD) in adolescence is a potent risk factor for major depressive disorder, yet its neurobiological mechanisms remain unclear. The glymphatic system, a brain-wide waste clearance pathway, has been implicated in psychiatric disorders but has not been investigated integratively in StD. This study aimed to characterize glymphatic function in adolescents with StD using a multimodal neuroimaging approach.

**Methods:** A total of 107 adolescents (71 with StD and 36 healthy controls) underwent multimodal MRI. Glymphatic function was assessed using three complementary metrics: the diffusion tensor imaging analysis along the perivascular space (DTI-ALPS) index for clearance capacity, BOLD-CSF coupling for neurovascular-driven circulation, and Hurst exponent for CSF flow dynamics. An integrated Mahalanobis Distance (MD) was calculated to quantify individual deviation from the healthy glymphatic profile.

**Results:** Compared with healthy controls, adolescents with StD exhibited significant reductions in the left DTI-ALPS index and global BOLD-CSF coupling (p < 0.05). No significant group differences were found in structural volumes of the choroid plexus or perivascular spaces. The integrated MD metric significantly differentiated StD from controls and demonstrated stronger associations with clinical severity than single biomarkers. Specifically, glymphatic dysfunction was significantly correlated with greater depressive symptom severity, sleep disturbances, and general psychopathology.

**Conclusions:** Adolescents with StD exhibit distinct, lateralized impairments in glymphatic clearance and neurovascular driving mechanisms. These findings provide early physiological evidence for a dimensional model of depression, suggesting that glymphatic dysfunction is present before diagnostic criteria for major depression are met. The multivariate MD metric offers a sensitive biomarker for tracking early pathophysiological changes and potential intervention targets.

## Introduction

### 1.1 StD in adolescent

Adolescence is a critical neurodevelopmental period marked by profound changes in brain structure and function, yet it is also a heightened vulnerability to the onset of mental illness. Among these, subthreshold depression (StD) represents a significant and often overlooked clinical challenge. StD refers to a condition where individuals exhibit clinically significant depressive symptoms that cause substantial functional impairment but do not meet full diagnostic criteria for major depressive disorder (MDD)(1). This condition is far from benign; longitudinal studies demonstrate that adolescents with StD face a significantly elevated risk of progressing to full-blown MDD in adulthood. A comprehensive meta-analysis revealed that individuals with StD had a nearly two-fold increased risk of developing MDD compared to non-depressed controls(2). It has also been found that adolescents with StD had a similar prognosis to those meeting criteria for major depression in terms of later depression and suicidal behaviors (3). It is crucial to understand the neurobiological underpinnings of StD for effective intervention in child development(4).

### 1.2 StD and neuroimaging

Neuroimaging studies have documented brain abnormalities in StD versus HC, encompassing changes in gray matter volume (5,6), alterations in white matter microstructure (5), resting-state functional abnormalities(7–9), and aberrant task-related activations during reward, emotional, and cognitive processes(10–12). Recent meta-analyses reveal convergent StD abnormalities in the salience network (insula, ACC)(13,14) and frontostriatal reward circuit(10), suggesting potential diagnostic(15,16) and therapeutic(17,18) biomarkers.

### 1.3 Glymphatic function assessment methods

The hypothesis of the glymphatic system has been reported and rapidly developed in recent years (19). It operates as an integrated Cerebrospinal Fluid (CSF) pathway comprising three interconnected components: CSF production, circulation dynamics, and waste clearance through perivascular channels (20,21). The choroid plexus (ChP) serves as the primary site for CSF production, residing within the brain’s ventricular system and continuously generating CSF to initiate the glymphatic circulation cycle (22,23). CSF circulation operates via neurovascular coupling (24), where neural slow waves induce cerebral blood volume fluctuations that generate compensatory CSF flow. A coupling index quantifies this process by measuring synchronization between blood-oxygen-level-dependent (BOLD) and CSF flow patterns, BOLD-CSF coupling, providing a window into circulation efficiency (25). Recent evidence suggests that this coupling is not merely a passive process but is actively modulated by the neuromodulatory system. Specifically, the integrity of the basal forebrain cholinergic system, particularly the nucleus basalis of Meynert (BF4), which provides the primary cholinergic innervation to the cortex, has been shown to be crucial for regulating BOLD-CSF coupling. The volume of the BF4 region correlates with both cortical cholinergic density and the strength of BOLD-CSF coupling, indicating that the degeneration of these neurons may impair glymphatic function through a neurovascular mechanism (26).

Beyond synchronization strength, the intrinsic temporal dynamics of CSF flow also offer critical insights into glymphatic function. The Hurst exponent (27,28) CSF signals, as a measure of the scale-free, fractal-like dynamics of CSF signals(25), characterizes the long-range correlations and predictability of CSF bottom-up inflow. Importantly, alterations in CSF dynamics captured by the Hurst exponent demonstrate convergent validity with other glymphatic measurements. Recent evidence shows that higher CSF-H values in Alzheimer’s disease pathology correlate not only with pathological biomarkers (CSF Aβ42 and tau) and the APOE ε4 genetic risk factor(29) but also parallel the impaired BOLD-CSF coupling observed in neurodegenerative conditions (30–32). This convergence suggests that CSF-H captures a distinct yet complementary aspect of glymphatic dysfunction that complement the coupling-based measures of neurovascular-driven CSF flux.

The final component involves waste removal through perivascular spaces (PVS). It is a fluid-filled channels surrounding blood vessels that facilitate metabolite clearance (33). The diffusion tensor imaging-based Analysis along the Perivascular Space (DTI-ALPS) index offers an indirect assessment of glymphatic clearance capacity by measuring water molecule diffusion along these spaces (34–36). To date, there is no gold standard metric for glymphatic function. Together, we hypothesize that these metrics form a comprehensive assessment of glymphatic health: BOLD-CSF coupling quantifies the neurovascular drive of CSF circulation, while CSF-H characterizes the resulting flow dynamics and efficiency.

### 1.4 Glymphatic system and mental disorder

Emerging evidence suggests an intriguing link between mental disorders and glymphatic system dysfunction, providing a compelling rationale for investigating this system in StD. Recent comprehensive reviews have documented correlations between the glymphatic system and neuropsychiatric and neurodegenerative conditions (37,38). In early psychosis, recent investigations have demonstrated that glymphatic clearance, as measured by BOLD-CSF coupling, is significantly reduced and delayed compared to healthy controls, with these alterations correlating with both cognitive decline and more severe psychotic symptoms (39). Similarly, in neurodegenerative language disorders, glymphatic dysfunction has been found to exhibit weaker global BOLD-CSF coupling and anterior-predominant DTI-ALPS impairment showing spatial overlap with regions of proteinopathy (40,41). However, neuroimaging studies of glymphatic function in StD are still lacking. Given that the glymphatic system serves as a fundamental brain maintenance function for metabolic waste clearance and homeostasis,its dysfunction represents a common pathway to neurodegeneration (42).

This study therefore aims to pioneeringly conduct a comprehensive investigation of glymphatic system function in adolescents with StD using a multimodal MR neuroimaging approach. We sought to clarify whether glymphatic dysfunction is present in this early stage of depression and to what extent it relates to transdiagnostic clinical features.

## Methods

### 2.1 Procedure

The Ethics Committee of Zhuhai Hospital of Guangdong Provincial Hospital of Chinese Medicine approved this study protocol (ZF2023-401-01) and was conducted in accordance with the ethical standards of the relevant national and institutional committees on human experimentation and with the Declaration of Helsinki as revised in 2013. Written informed consent was obtained from all participants (and from their legal guardians for participants under 18 years of age) prior to their inclusion in the study. Recruitment took place between April 2024 and June 2025. Potential participants were first screened using the Self-Rating Depression Scale (SDS). Those who met the initial criteria were invited for a comprehensive clinical assessment conducted by experienced physicians. This assessment included a structured clinical interview based on the Diagnostic and Statistical Manual of Mental Disorders, Fifth Edition (DSM-5) criteria to finalize their group assignment (StD or HC group). During the interview, participants also completed a battery of self-report questionnaires. Within one week of the clinical assessment, eligible participants underwent a multimodal magnetic resonance imaging (MRI) scan. Participants were compensated for their time upon completion of the study.

### 2.2 Clinical assessments

Self-Rating Depression Scale (SDS) (43) is a 20-item self-report questionnaire used to screen depressive symptoms. Each item is rated on a 4-point scale. The raw total score is multiplied by 1.25 to obtain a standard score. A standard score ≥ 53 is considered indicative of clinically significant depressive symptoms and was used as a screening criterion in this study.

Patient Health Questionnaire-9 (PHQ-9) (44) is a 9-item self-report tool designed according to DSM-5 diagnostic criteria for depression to assess the severity of depressive symptoms. Each item is scored from 0 (“Not at all”) to 3 (“Nearly every day”), with a total score ranging from 0 to 27. Higher scores indicate greater depression severity.

Symptom Checklist-90 (SCL-90) (45) is a 90-item self-report inventory used to evaluate a broad range of psychological problems and psychopathology. It covers nine primary symptom dimensions and provides three global indices of distress. Higher scores reflect greater levels of psychological distress.

Insomnia Severity Index (ISI) (46) is a 7-item self-report questionnaire that assesses nature, severity, and impact of insomnia. Each item is rated on a 5-point scale from 0 to 4, with a total score ranging from 0 to 28. Higher scores indicate more severe insomnia.

Pittsburgh Sleep Quality Index (PSQI) (47) is a 19-item self-report questionnaire that assesses sleep quality and disturbances over the past month. It evaluates seven components: subjective sleep quality, sleep latency, sleep duration, habitual sleep efficiency, sleep disturbances, use of sleeping medication, and daytime dysfunction. A global score > 5 is generally indicative of poor sleep quality.

### 2.3 Participant inclusion and exclusion criteria

Adolescents aged 15–22 years were recruited between April 2024 and June 2025. All participants were divided into two groups: the subthreshold depression (StD) group and the healthy control (HC) group.

#### Inclusion criteria for the StD group

(1) Self-Rating Depression Scale (SDS) score ≥ 53; (2) presence of ≥ 2 depressive symptoms persisting over the past two weeks but not meeting the Diagnostic and Statistical Manual of Mental Disorders, Fifth Edition (DSM-5) criteria for major depressive disorder (MDD), which requires ≥ 5 symptoms; (3) right-handedness; (4) age 15–22 years; (5) no history of traumatic brain injury, and normal or corrected-to-normal vision.

#### Inclusion criteria for the HC group

(1) SDS score < 53; (2) right-handedness; (3) age 15–22 years; (4) no history of traumatic brain injury, and normal or corrected-to-normal vision; (5) no family history of psychiatric disorders.

#### Exclusion criteria for all participants

(1) history of substance abuse; (2) severe neurological disorders (e.g., post-concussion syndrome, neuroinflammatory diseases) or related treatment history (e.g., intracranial space-occupying lesions, chronic neurological diseases); (3) contraindications for magnetic resonance imaging (MRI) examination (e.g., metallic implants, claustrophobia); (4) poor MRI image quality.

Following the initial screening with the SDS, all participants underwent clinical interviews. The assessments were conducted by experienced physicians from the Department of Psychology at Zhuhai Hospital of Guangdong Provincial Hospital of Chinese Medicine. The diagnostic evaluation was based on the DSM-5 criteria for depressive disorders, including symptom manifestation and duration criteria. For the StD group, the assessment confirmed that participants did not meet the full diagnostic criteria for MDD. For the HC group, the assessment ensured no history of psychiatric disorders.

### 2.3 Image acquisition

All images were acquired on a GE DISCOVERY MR750w 3.0T scanner using a standard head coil.

High-resolution T1-weighted anatomical images were acquired using a magnetization-prepared rapid gradient-echo (MPRAGE) sequence with the following parameters: Repetition Time (TR) = 8.488 ms; Echo Time (TE) = 3.248 ms; Inversion Time (TI) = 450 ms; flip angle = 12°; matrix size = 256 × 256; voxel size = 1×1×1 mm³.

Resting state functional images were acquired using a gradient-echo echo-planar imaging (GRE-EPI) sequence with the following parameters: repetition time (TR) = 2000 ms; echo time (TE) = 30 ms, flip angle = 90°, slice thickness = 4 mm, spacing between slices = 4 mm, field of view (FOV) = 224×224 mm², matrix size = 64×64, voxel size = 3.5×3.5×4.0 mm³. A total of 37 axial slices were acquired, and the scan duration was 490 seconds, resulting in 245 volumes.

Diffusion-weighted images were acquired using a spin-echo echo-planar imaging (SE-EPI) sequence with following acquisition parameters: Repetition Time (TR) = 11500 ms; Echo Time (TE) = 75 ms, flip angle = 90°, slice thickness = 2mm, spacing between slices = 3mm, field of view (FOV) = 256×256 mm³, matrix size = 128×128, reconstruction matrix = 256×256, voxel size = 2×2×2 mm³. A total of 54 axial slices were acquired. Diffusion gradients were applied along 32 non-collinear directions with a b-value of 1000 s/mm², in addition to 1 non-diffusion-weighted (b=0) images.

### 2.4 BOLD-CSF coupling

whole-brain gray matter BOLD time series and CSF signals were extracted from functional MRI data for each participant. Both signals underwent preprocessing with DPABI software^54^, including removal of the first 10 volumes, slice timing correction, motion correction, quadratic detrending, and bandpass filtering (0.01-0.1 Hz). For CSF signal extraction, T1 images were registered to standard space via SPM12 (48) to create CSF masks, with signals extracted from the first valid CSF mask slice in inferior-to-superior order. Each mask was validated against individual T1 images to ensure segmentation accuracy. We conducted visual assessment of CSF mask segmentation quality for each participant, excluding participants whose segmentation failed due to incomplete cerebellar coverage in the field of view. For BOLD signal processing, white matter and CSF nuisance signals were regressed, followed by spatial normalization to MNI space using DARTEL and smoothing with a 6mm FWHM Gaussian kernel. Final BOLD signals were extracted from gray matter regions defined by the Brainnetome Atlas (BNA246).

### 2.5 Hurst exponent of CSF

Initial preprocessing was performed on the 4D fMRI time series for each subject. A mean functional image was computed by averaging the entire series. Subsequently, this mean image was segmented into gray matter (GM), white matter (WM), and cerebrospinal fluid (CSF) probability maps using the Segmentation tool within the SPM12 software package.

To accurately define the CSF space, the resulting CSF probability map was thresholded. Voxels with a probability value of 0.9 or greater were classified as CSF, while all other voxels were excluded. This process generated a binarized CSF mask for each individual subject.

The subject-specific binarized CSF mask was applied to the original 4D fMRI data to isolate signals originating from the CSF space. To specifically capture the dynamics of CSF inflow, a unique time-series extraction method was employed. For each point (volume) in the fMRI scan, we identified the superior-most axial slice (the slice with the highest Z-axis coordinate) containing CSF voxels. The mean signal intensity of all non-zero voxels within this specific slice was then calculated. This procedure yielded a single time series for each subject, representing the dynamic fluctuations of CSF inflow over time.

To assess the long-term memory and self-similarity of the CSF inflow signal, the Hurst exponent was calculated for each subject’s extracted CSF time series. The classic Rescaled Range (R/S) analysis was used for this calculation (49,50).

The specific steps of the R/S analysis were as follows:

1. The CSF time series of length N was divided into multiple non-overlapping sub-series of varying lengths “n” (ranging from 10 to N/2).
2. For each sub-series, the range “R” (difference between the maximum and minimum of the cumulative deviation series) and the standard deviation “S” were calculated.
3. The rescaled range, “R/S”, was then computed.
4. The “R/S” values were averaged across all sub-series of the same length “n”.
5. Finally, a linear regression was performed on a log-log plot with the logarithm of the sub-series length as the independent variable and the logarithm of the averaged “R/S” value as the dependent variable. The slope of this regression line is the Hurst exponent.

The Hurst exponent ranges from 0 to 1. A value of H > 0.5 indicates a persistent or long-term memory process, H < 0.5 suggests an anti-persistent process, and H = 0.5 corresponds to a completely random series.

### 2.6 DTI-ALPS index

We calculated ALPS for each participant using diffusion MRI data processed entirely with FSL tools^57^. Raw diffusion data underwent preprocessing including brain extraction using BET, motion and eddy current correction via eddy correct, and susceptibility distortion correction using top up with reversed phase-encoding acquisitions. DTI fitting was performed using dti-fit to generate diffusivity parameter maps (FA, MD, AD, RD).

All DTI maps were registered to the JHU-ICBM-FA-1mm template using FLIRT for linear registration followed by FNIRT for nonlinear alignment. Four regions of interest were defined from the JHU white matter atlas: bilateral superior corona radiata (projection fibers) at coordinates L_SCR (53,47,40) and R_SCR (25,47,40), and bilateral superior longitudinal fasciculus (association fibers) at L_SLF (59,47,40) and R_SLF (19,47,40).

Directional diffusivity values (Dxx, Dyy, Dzz) were extracted from these ROIs for ALPS calculation. The ALPS index was computed as the bilateral mean ratio^58^:

ALPS = (mean(Dxxproj + Dxxassoc))/(mean(Dyyproj + Dzzassoc)), where Dxxproj/assoc represents x-axis diffusivity in projection/association fibers, Dyyproj represents y-axis diffusivity in projection fibers, and Dzzassoc represents z-axis diffusivity in association fibers.

### 2.7 Chp volume

Volume Choroid plexus was segmented for each participant from structural MRI^51^. Specifically, T1-weighted images were registered to ICBM-MNI 152 standard space using ANTs (51)to reduce morphological variability. Choroid plexus segmentation was performed using a deep learning approach that demonstrated superior performance (Dice coefficient: 0.72, Hausdorff distance: 1.97mm) compared to atlas-based methods^52^. The model was trained on 64×64×64 voxel patches with data augmentation, and segmentation results were transformed back to native space for volume extraction. Raters independently verified segmentation quality across all participants. Total Intracranial Volume (TIV) was calculated using SPM12^53^ by summing gray matter, white matter, and cerebrospinal fluid volumes, selected for its demonstrated consistency and low variability. Normalized choroid plexus volume was expressed as the percentage ratio of absolute choroid plexus volume to TIV.

### 2.8 PVS volume

PVS volume was extracted using the SHIVA(52) (https://github.com/pboutinaud/SHIVA_PVS) pipeline, which leverages an autoencoder and U-Net architecture (53) for medical image segmentation. The preprocessing workflow began with data acquisition, where T1-weighted images and corresponding brain masks were loaded using a “datagrabber” node. Subsequently, all images were resampled to 1mm isotropic voxels and reoriented to a consistent orientation using the “conform” node to ensure dimensional and orientational uniformity across subjects while preserving the native space of each scan. Intensity normalization was then performed within the brain-masked region to standardize signal intensities across subjects. A precise brain mask was generated for each subject using the “proper_brain_mask” node. To remove non-brain tissue such as the neck and shoulders, the images were cropped based on the bounding box of the brain mask. Finally, to ensure patient privacy, the images were de-faced using the “defacing_img1” node.

Following preprocessing, perivascular spaces (PVS) were segmented from the T1-weighted images using a pre-trained U-Net model. This model was trained on a large, expertly curated dataset of manual PVS segmentations to ensure high accuracy.

The PVS segmentation maps were extracted from the model’s output for further analysis. For quality control and visualization purposes, the segmented PVS were overlaid on the original T1-weighted images. Quantitative analysis was performed to characterize the PVS burden. A connected-component analysis was applied to the PVS segmentations using the “cluster_labelling_pvs” node to identify and count individual PVS. From this, the total volume and number of PVS were calculated for each subject.

Multiple quality control (QC) steps were integrated throughout the workflow to ensure the reliability of the outputs at each stage. The “qc_crop_box” node was used to verify the accuracy of the image cropping. The alignment of the brain mask with the T1-weighted image was validated using the “qc_overlay_brainmask” node. Various image quality metrics were computed using the “qc_metrics” node. Finally, a comprehensive QC report was generated for each subject using the “summary_report” node, summarizing the results of all checks.

### 2.9 BF4

The volumetric quantification of the BF4 was performed utilizing the SPM Anatomy Toolbox^76,82^ implementation of stereotaxic probabilistic maps of magnocellular cell groups in the human basal forebrain^83^. Specifically, we extracted volumetric data corresponding to the Ch4 cell group, which anatomically represents the BF4 region. Consistent with established protocols in previous investigations^84^, we utilized absolute volumetric measurements rather than normalized values for this analysis, given the relatively small volume of the BF4 structure and the potential introduction of normalization-related variance in small subcortical nuclei.

### 2.10 MD

We established a multivariate reference framework using the healthy control (HC) group to calculate individualized Mahalanobis distances (54) for each participant(55).

The HC group served as the normative population from which we derived the reference mean vector (μ) and covariance matrix (Σ) across six lymphatic biomarkers. For each individual participant (both StD patients and HC subjects), we computed Individual MD for using:

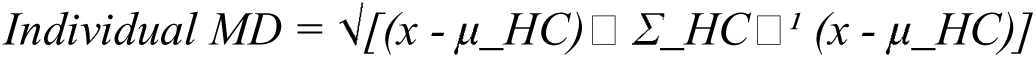

where x represents the individual’s biomarker profile, μ_HC is the HC group mean vector, and Σ_HC□^1^ is the inverse covariance matrix derived from the HC population. This approach quantifies each person’s multivariate deviation from the healthy reference standard, providing a standardized measure of glymphatic system dysfunction. Higher individual MD values indicate greater deviation from normal healthy patterns, enabling direct comparison of dysfunction severity across all participants regardless of their diagnostic group.

### 2.11 Statistical analysis

To assess the first hypothesis, we conducted analysis of covariance (ANCOVA) to compare glymphatic biomarkers between the StD and HC groups across six biomarker series, with age and sex as covariates (p < 0.05). Within each biomarker series, we selected the marker with the highest statistical power for subsequent clinical correlation analyses and inclusion in the Mahalanobis distance model.

To test Hypothesis 2, we performed partial correlation analyses between the six selected biomarkers from Hypothesis 1 testing and transdiagnostic behavioral traits, controlling for age and sex as covariates (p < 0.05).

To test Hypothesis 3, we established a reference baseline using the six biomarkers from the HC group to construct the Mahalanobis distance framework. Individual Mahalanobis distances were then calculated for each StD subject relative to this HC reference. To evaluate the discriminative power of individualized glymphatic function distance for StD symptoms at both categorical and continuous levels, we conducted group comparisons between StD and HC groups using ANCOVA with age and sex as covariates (p < 0.05), followed by partial correlation analyses between individual Mahalanobis distance values and depression-related clinical measures, controlling for age and sex (p < 0.05).

## Results

### 3.1 Participants characteristics

A total of 107 adolescents participated in this study, including 71 individuals with StD and 36 HC. Demographic and clinical characteristics of the two groups are presented in Table 1. The groups did not significantly differ in age (StD: M = 17.11, SD = 1.35; HC: M = 17.17, SD = 2.09) nor the proportion of only children. However, the StD had a higher propotion of females (67.6%) compared to the HC group (47.2%).

**Table 1.** Demographic Characteristics and Baseline Clinical Measures.

| Characteristic | Depression Group<br>(n = 71) | Healthy<br>Control<br>(n = 36) | Statistic |
| --- | --- | --- | --- |
| Demographics |  |  |  |
| Age, M (SD) | 17.11 (1.35) | 17.17 (2.09) | $t(105) = -0.16, p = 0.872$ |
| Female, n (%) | 48 (67.6) | 17 (47.2) | $\chi^2(1) = 3.35, p = 0.067$ |
| Only child, n (%) | 10 (14.1) | 4 (11.1) | $\chi^2(1) = 0.02, p = 0.898$ |
| Single-parent family, n<br>(%) | 9 (12.7) | 2 (5.6) | $\chi^2(1) = 0.65, p = 0.418$ |
| Clinical Measures |  |  |  |
| SDS, M (SD) | 67.92 (7.82) | 43.50 (5.78) | $t(105) = 16.57, p < .001$ |
| PHQ-9, M (SD) | 12.16 (5.89) | 1.81 (1.37) | $t(71) = 10.27, p < .001$ |
| SCL-90 Total, M (SD) | 226.70 (71.90) | 120.86 (16.98) | $t(105) = 8.69, p < .001$ |
| SCL-90 Mean, M (SD) | 2.52 (0.80) | 1.34 (0.19) | $t(105) = 8.70, p < .001$ |
| ISI, M (SD) | 9.94 (6.34) | 3.31 (3.66) | $t(98) = 5.68, p < .001$ |
| PSQI, M (SD) | 9.15 (3.53) | 5.26 (2.80) | $t(98) = 5.64, p < .001$ |

*Note.* SDS = Self-Rating Depression Scale; PHQ-9 = Patient Health Questionnaire-9; SCL-90 = Symptom Checklist-90; ISI = Insomnia Severity Index; PSQI = Pittsburgh Sleep Quality Index. M = mean; SD = standard deviation.

The StD group showed significantly higher scores on all clinical measures compared to HC group. Specifically, StD group exhibited elevated depressive symptoms as measured by Self-rating Depression Scale (SDS: M = 67.92, SD = 7.82 vs. M = 43.50, SD = 5.78) and the Patient Health Questionnaire-9 (PHQ-9: M = 12.16, SD = 5.89 vs. M = 1.81, SD = 1.37). The StD group also demonstrated higher levels of general psychopathology on the Symptom Checklist-90 (SCL-90 total: M = 226.70, SD = 71.90 vs. M = 120.86, SD = 16.98) and more severe sleep disturbances as indicated by the Insomnia Severity Index (ISI: M = 9.94, SD = 6.34 vs. M = 3.31, SD = 3.66) and Pittsburgh Sleep Quality Index (PSQI: M = 9.15, SD = 3.53 vs. M = 5.26, SD = 2.80). These differences confirmed successful group classification and highlighted the clinical burden associated with subthreshold depressive symptoms.

### 3.2 Intercorrelations among glymphatic biomarkers

We investigated intercorrelations among the glymphatic function indices before examining group differences adjusted for age and gender. The partial correlation matrix is presented in Table S1 and visualized in Figure 3B-C.

**Figure 1.**
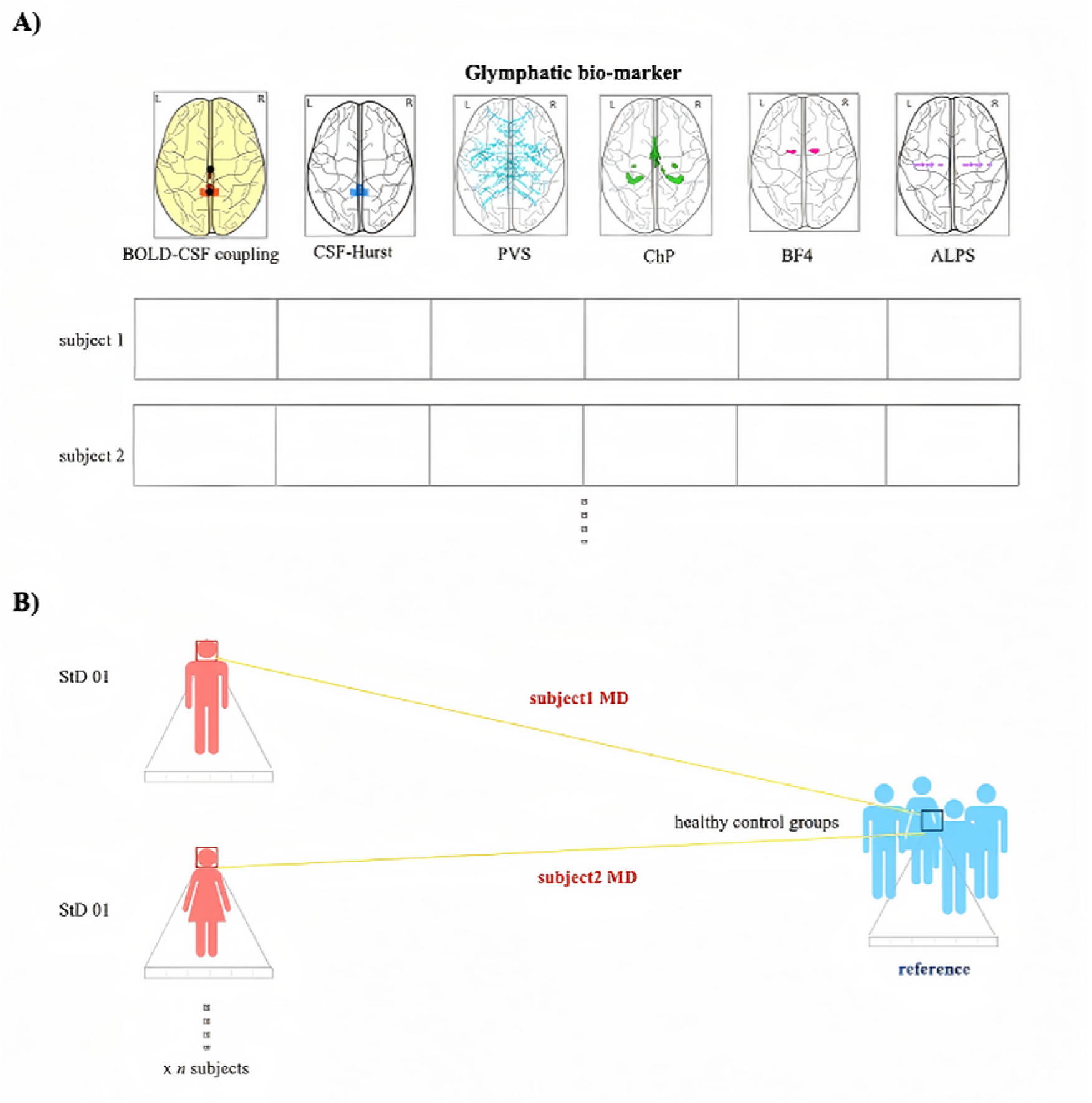
Conceptual Framework of Multivariate Mahalanobis Distance (MD) for Assessing Glymphatic Dysfunction. This figure illustrates the integration of multiple neuroimaging biomarkers to quantify individual glymphatic health. A. A reference “normal” space (represented by the ellipsoid) is constructed based on the multidimensional glymphatic profile of the healthy control (HC) group. The center point represents the mean vector of the healthy population. B. The Mahalanobis distance (MD) is calculated for each participant, including those with subthreshold depression (StD), relative to this healthy reference center. A larger MD value indicates a greater deviation from the healthy baseline, serving as a standardized, composite metric of glymphatic system dysfunction.

**Figure 2.**
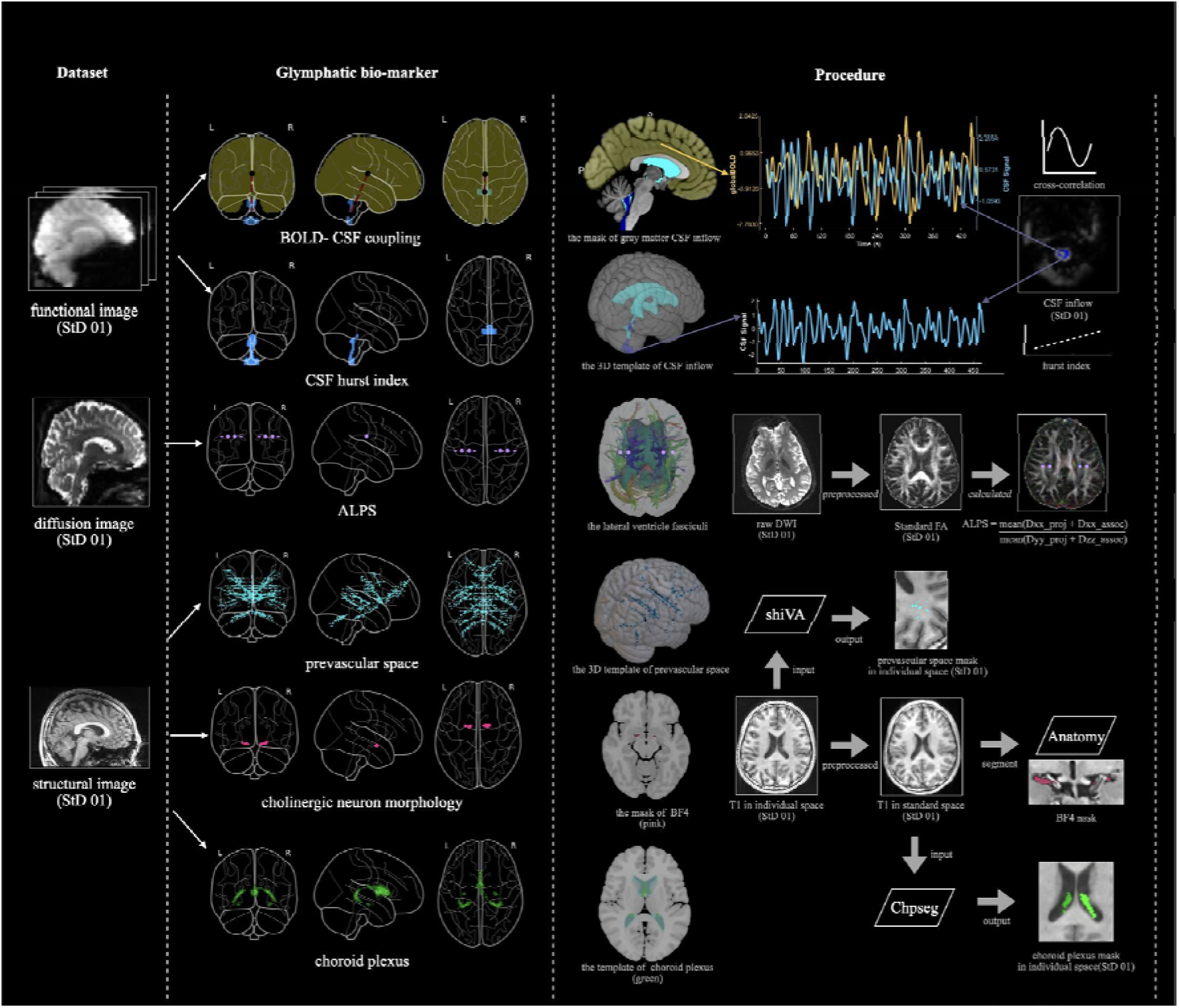
Multimodal Neuroimaging Approaches for Glymphatic System Assessment. Comprehensive evaluation of glymphatic system components using multimodal MRI. (A) BOLD-CSF Coupling: Illustration of the extraction of global gray matter BOLD signals and cerebrospinal fluid (CSF) signals from the fourth ventricle, followed by the calculation of their synchronization to quantify neurovascular-driven CSF circulation. (B) CSF Dynamics: Application of Hurst exponent analysis to the CSF inflow time series to characterize the temporal dynamics and long-range correlations of fluid flow. (C) DTI-ALPS Index: Placement of regions of interest (ROIs) in projection and association fibers at the level of the lateral ventricle body on diffusion tensor imaging (DTI) maps to calculate diffusivity along perivascular spaces. (D) Structural Segmentation: Automated segmentation and volumetric quantification of the choroid plexus (ChP), perivascular spaces (PVS), and the basal forebrain cholinergic nucleus (BF4).

**Figure 3.**
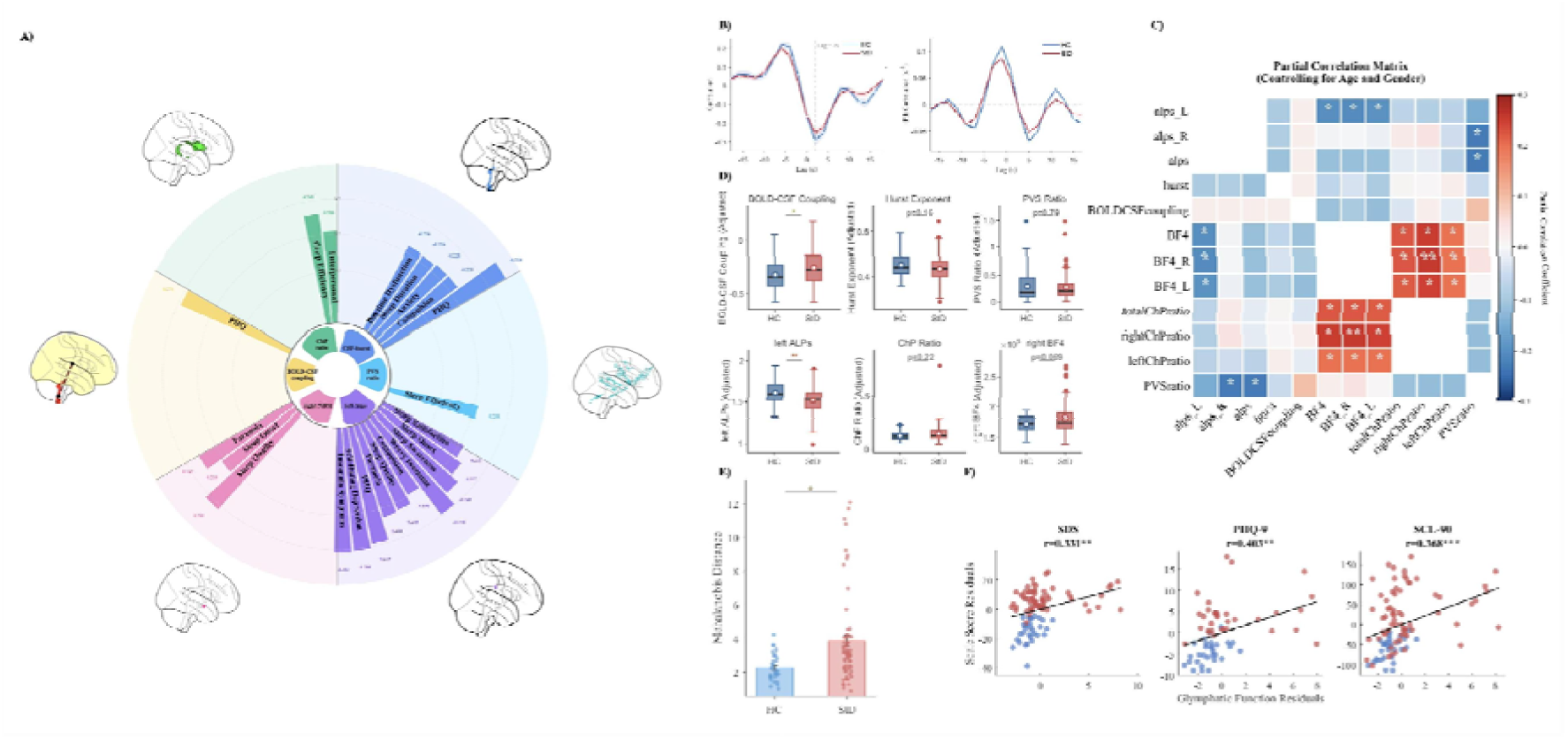
Glymphatic System Dysfunction in Adolescents with Subthreshold Depression (StD) and Clinical Associations. (A) Group Comparisons: Violin plots depict significant differences in key glymphatic metrics between the StD group (red) and Healthy Controls (HC, blue). The StD group exhibits significantly reduced Left DTI-ALPS index and BOLD-CSF coupling, along with a significantly increased integrated Mahalanobis Distance (MD) compared to HC (p < 0.05). (B-C) Biomarker Intercorrelations: Heatmaps show the partial correlation matrix among the six glymphatic biomarkers, adjusting for age and sex, revealing intrinsic relationships between different glymphatic components. (D-F) Clinical Correlations: Scatter plots demonstrate significant associations between glymphatic function and transdiagnostic clinical symptoms. Lower Left ALPS values correlate with higher depression severity (SDS, PHQ-9) and sleep disturbances (ISI, PSQI). Higher MD values are associated with greater depressive symptoms (PHQ-9) and general psychopathology (SCL-90), indicating that more severe glymphatic dysfunction tracks with higher clinical burden. Error bands represent 95% confidence intervals. Abbreviations: StD, subthreshold depression; HC, healthy control; ALPS, analysis along the perivascular space; BOLD, blood-oxygen-level-dependent; CSF, cerebrospinal fluid; ChP, choroid plexus; PVS, perivascular space; BF4, basal forebrain region 4.

### 3.3 Group difference of glymphatic measures

We first evaluated if individuals with StD would exhibit glymphatic dysfunction compared to healthy controls, we conducted a series of ANCOVAs for each glymphatic biomarker, controlling age and gender. Bilateral ALPS showed a trend toward significance (F(_1,103_) = 3.77, p = .055), which appeared to be mainly driven by left hemisphere effect. Right hemisphere ALPS showed no significant group difference (F(_1,103_)) = 0.24, p = .623), suggesting asymmetry in glymphatic alterations associated with StD. Age and gender effect were not significant for any ALPS measure. BOLD-CSF coupling demonstrated a significant group effect (F (_1,103_) = 4.14, p = 0.044, StD > HC). This indicated decreased synchronization between BOLD signal fluctuations and CSF dynamics in StD group. Neither age (F = 1.83, p = .179) nor gender (F = 0.59, p = .445) showed significant effects. Hurst exponent of CSF inflow effect did not show significant group differences (F(_1,103_) = 1.97, p = .164). BF4 volume approached but did not reach statistical significance for any measure(Bilateral: F(_1,103_) = 3.17, p = .078; Right: F(_1,103_) = 3.39, p = .069; Left: F(_1,103_) = 2.85, p = .094). These marginally significant findings suggest a potential reduction in the basal forebrain cholinergic system in StD. ChP ratio showed no significant group difference (F(_1,103_) = 1.93, p = .168). PVS also showed no significant group difference (F(_1,103_) = 0.07, p = .788).

### 3.4 Associations between glymphatic function and transdiagnostic clinical symptoms

We investigated if glymphatic function would be associated with key transdiagnostic clinical dimensions using partial correlation analyses controlling age and gender.

Left ALPS emerged as the most broadly associated glymphatic measures with clinical dimensions. Lower left ALPS values were significantly correlated with core depressive symptoms (higher SDS total scores: r = −.29, p = .004; higher PHQ-9 total scores: r = −.28, p = .022), sleep disturbance (higher ISI: r = −.29, p = .005; higher PSQI: r = −.22, p = .033; greater ISI-5: r = −.28, p = .006; increased ISI-6: r = −.25, p = .015; ISI-1: r = −.23, p = .026; lower ISI-4: r = −.21, p = .039), and general psychopathology (higher SCL-90 - obsessive-compulsive: r = −.20, p = .045; higher SCL-90 - paranoid ideation: r = −.22, p = .024). The particularly strong associations with sleep measures are noteworthy given the critical role of sleep in glymphatic clearance.

Hurst exponent showed selective associations with mood and anxiety symptoms (higher PHQ-9: r = −.32, p = .007; higher SCL-90 - obsessive-compulsive: r = −.22, p = .025; higher SCL-90 - anxiety: r = −.22, p = .026) as well as sleep-related features (shorter PSQI – sleep duration: r = −.22, p = .036; greater PSQI – daytime dysfunction: r = −.22, p = .035). Note that results of Hurst exponent are partially paralleled with part of left ALPS’s results.

Interestingly, While total Chp ratio showed two associations with interpersonal and sleep dimensions (higher SCL-90 – interpersonal sensitivity: r = .21, p = .034; higher PSQI – sleep efficiency: r = .25, p = .014), another positive association was discovered in PVS ratio for the same dimension (higher PSQI – sleep efficiency: r = .21, p = .045) (Table 3A).

### 3.5 Mahalanobis Distance as an Integrated Glymphatic Function Metric

We further explored whether integrating multiple glymphatic biomarkers using Mahalanobis distance (MD) would provide a better performance in discriminating between StD and HC and in predicting clinical symptoms. Average MD values significantly differentiated from the StD (StD: M =4.07; HC: M = 2.41, ANCOVA F = 6.17, p = .01). The correlation between MD and PHQ-9 was stronger (r = .403, p= .001, n = 63) than the strongest single biomarker correlation (left ALPS: r = -.28) and was also correlated to both general psychopathology (total SCL-90: r = .37, p = .0002, n = 95)and self-assessment score (total SDS: r = .33, p = .0011, n = 95), suggesting a superior capability of catching variability of depression severity.

## Discussion

### 4.1 Main findings

This study represents the pioneering comprehensive, multimodal MR investigation of glymphatic system function in adolescents with StD, a clinically significant population that falls below the arbitrary thresholds of current categorical systems yet experiences substantial functional impairment (56).

The first important findings are significant left-hemispheric reductions in the DTI-ALPS index accompanied by decreased BOLD-CSF coupling in StD participants compared to healthy controls.To our knowledge, this is the first study to specifically identify a left-hemispheric reduction of the DTI-ALPS index in a population with depressive symptoms, particularly during the critical neurodevelopmental window of adolescence and at the subthreshold stage. The left-lateralized deficit strongly suggests mechanisms tied to the well-documented hemispheric asymmetries in mood regulation (57). The left hemisphere is critical for approach-related emotions and positive affect (58), functions characteristically impaired in depression. Left-lateralized glymphatic failure could therefore reflect either the cause or a consequence of reduced metabolic activity in these vital affect-regulatory circuits (59). This hypothesis is indirectly supported by evidence of impaired resting-state functional connectivity within the cognitive control network (CCN) in StD individuals, a network crucial for regulating self-referential thought and known to be dysfunctional in depression (60). A localized failure in glymphatic clearance could therefore be both a cause and a consequence of such network-level dysregulation (61).

The lateralization pattern reveals a notable asymmetry in glymphatic dysfunction. In our analysis, the fact that bilateral ALPS was only trend-significant while the left-hemisphere effect was robust. It reveals that averaging across hemisphere may dilute focal, lateralized pathology particularly relevant to early-stage depression. This finding is particularly noteworthy given recent reports of hemispheric asymmetry in glymphatic dysfunction in Parkinson’s disease (62) suggesting that lateralized glymphatic impairment may be a transdiagnostic dimension of neuropsychiatric disorders.

### 4.2 Reconciling divergent findings

#### 4.2.1 Emerging controversies in DTI-ALPS methodology

We acknowledged that a recent study by Zhang et al. (63) reported no significant DTI-ALPS alterations either bilateral or lateralized in MDD adolescents. Part of our results appears to contrast with above null findings. However, careful examination reveals that these studies may in fact be complementary, highlighting important considerations about disease stage, methodological sensitivity, and the interpretation of glymphatic imaging findings in psychiatric populations.

Recent methodological critiques have raised significant concerns about DTI-ALPS as a standalone marker of glymphatic function. Specifically, DTI-ALPS measures are substantially confounded by white matter geometry – including crossing fibers, axonal undulations, and dispersion – factors that may obscure genuine perivascular diffusion signals (64–66). The influential review by Taoka et al. (35) explicitly cautioned that decreased ALPS-index should be described only as “decrease ALPS-index” rather than equalizing with “glymphatic dysfunction”.

#### 4.2.2 The critical advantage of multimodal assessment

In contrast to reliance on DTI-ALPS alone, our multimodal approach integrates functional dynamics and structural metrics. As these metrics showed near-zero intercorrelations in our data, they capture complementary dimensions and integrated multimodal deviation of glymphatic function. This methodological triangulation enhances sensitivity to detect subtle glymphatic alterations that may be masked by structural confounds in single-modality assessments, particularly when facing methodological uncertainties inherent to each individual measures.

### 4.3 Neurobiological mechanisms and clinical associations

#### 4.3.1 BOLD-CSF coupling and neurovascular dynamics

The BOLD-CSF coupling result resonates with previous work characterizing glymphatic dysfunction in neuropsychiatric disorders and highlights adolescence as a critical period for such dysfunctions to emerge. (As this metric showed a near-zero correlation with ALPS indices, it captures a distinct dimension of glymphatic function related to the dynamic vascular-CSF interactions that drive fluid movement.) The decreased coupling intensity may represent a failed compensatory response to the clearance deficits indexed by ALPS, or it could reflect the neurovascular uncoupling characteristic of depression (67). Notably, these findings parallel structural alterations reported in prior neuroimaging studies showing reduced gray matter volume in the orbitofrontal cortex and temporal gyrus in both StD and MDD populations (68) reinforcing the notion that glymphatic dysfunction is intimately connected to the broader neuropathology of the depression spectrum.

#### 4.3.2 Transdiagnostic clinical associations

Our dimensional analyses revealed pervasive associations between glymphatic biomarkers and a broad range of transdiagnostic clinical features including depressive symptom severity, sleep disturbances, and general psychopathology, especially left ALPS, and Hurst exponent of CSF inflow. This pattern supports the conceptualization of glymphatic dysfunction as a shared biological vulnerability factor that transcends traditional diagnostic boundaries. This transdiagnostic pattern supports dimensional models of psychopathology, where glymphatic dysfunction may function as a shared vulnerability factor. The presence of such clear biological markers at the subthreshold stage suggests that the pathophysiology of depressive disorders develops along a continuum.

#### 4.3.3 Sleep, glymphatic function, and disease stage

The relationship between sleep and glymphatic function warrants particular attention given discrepancies that finding of no correlation between subjective sleep quality (PSQI) scores. Several factors may explain this inconsistency. First, PSQI captures subjective sleep satisfaction rather than objective sleep architecture. Glymphatic function may be more closely related to objective parameters. Second, our findings of correlations between sleep disturbance and left ALPS in the StD cohort (Table S3) suggest that sleep-glymphatic relationships may be intact during early disease stages. In established MDD, however, other factors such as disease comorbidity and chronicity may obscure this association. Third, Depression itself causes negative cognitive biases that may inflate subjective sleep complaints independent of acute change of sleep architecture. This dissociation between subjective perception and objective physiology could explain why PSQI show no relationship with ALPS in MDD group while we observed associations in StD, where depressive cognitive distortions may be less pronounced.

### 4.4 A dimensional framework: MD as multivariate integration

Currently, there are no gold standard measurements for glymphatic function non-invasively. To address this central challenge, several multivariate approaches have been implemented, leading to recent advances in psychiatric neuroimaging. The MD method offers several advantages that complement other multivariate techniques. For example, it provides a powerful and computationally efficient metric at the individual level, extending the group-level comparisons offered by methods like Hotelling’s T-squared test (69,70). Another key strength of the MD metric is that it inherently accounts for the covariance structure among biomarkers, a critical feature for interpreting multimodal imaging data where measures are often intercorrelated (71). This makes it a particularly suitable and versatile method for neuroimaging. Data-driven methods, such as the radiomics-based machine learning strategy to discriminate between StD and MDD with high accuracy, highlight the power of synthesizing multi-scale features (72). The MD metric aligns with this data-driven paradigm with greater interpretability. The MD framework allows for the explicit examination of each biomarker’s contribution to an individual’s overall deviation score, providing potential mechanistic insights (55). Furthermore, its robust statistical foundation is well-suited for the sample sizes typically available in clinical studies, for which complex machine learning models may require larger datasets.

Our MD results provide a clinically intuitive measure of an individual’s overall deviation from a healthy baseline and show better discriminating power between StD and HC than any single glymphatic measure. It also exhibits a stronger association with symptoms of depression. This superior performance suggests that the MD metric optimally captures complementary information from each glymphatic measure. By providing a comprehensive, integrated assessment framework, the results validate our conceptual model of multidimensional impairment and offer a practical path toward clinical translation. Future clinical trials could stratify participants by glymphatic MD rather than diagnostic categories, testing whether high-MD individuals selectively respond to clearance-enhancing interventions, while low-MD individuals require alternative approaches.

### 4.5 Implications and perspectives for dimensional models of depression

For over a decade, systematic reviews of clinical and epidemiological data have argued that depression is better understood as a dimensional spectrum rather than a discrete category, challenging the arbitrary symptom counts used in current categorical diagnostic systems (73). However, many of these debates have lacked direct neurobiological evidence from the subthreshold population itself. Our dimensional analyses provide direct evidence for the very beginning of this depressive spectrum. The continuous correlation between the integrated glymphatic metric and clinical symptom severity across both HC and StD participants demonstrates a gradient of dysfunction rather than a categorical shift. This finding suggests that the neurobiological underpinnings of depression develop along a continuum, starting from a state of health. While the absence of an MDD group prevents a full characterization of the full spectrum, our study provides a unique and vital contribution by capturing the neurobiology of its earliest stages, establishing a critical foundation for the spectrum model of depression. Examine whether glymphatic dysfunction dimensions cut across traditional boundaries is needed in future work to be suggested.

#### 4.5.1 Early intervention and prevention

The demonstration of measurable glymphatic dysfunction in StD identifies a critical window for preventive intervention. If glymphatic impairment represents an early, potentially reversible pathophysiological process, then interventions targeting clearance mechanisms during the StD phase might prevent progression into MDD. Longitudinal studies tracking glymphatic function from StD through potential MDD onset are essential to test this preventive paradigm.

#### 4.5.2 Biomarker-guided treatment stratification

The integration of glymphatic metrics via MD enables a precision psychiatry approach. Rather than treating all StD individuals identically, clinicians could stratify patients based on glymphatic dysfunction severity. This stratified approach could improve treatment response rates and reduce trial-and-error prescribing. Addressing the substantial non-response problem in adolescent depression treatment.

### 4.6 Limitations

Several limitations warrant consideration. The primary limitation is our cross-sectional design, which precludes causal inferences regarding the temporal relationship between glymphatic dysfunction and the development of depressive symptoms. Longitudinal studies following StD individuals over time are essential to determine whether glymphatic impairment precedes symptom progression, represents a consequence of early pathophysiological changes, or both. Such studies could also clarify whether early intervention targeting glymphatic function (e.g., sleep optimization, exercise, or pharmacological enhancement of CSF dynamics) can prevent progression to MDD. The relatively modest sample size may have limited statistical power to detect smaller effects or examine subgroup differences. The sex distribution showed an overrepresentation of females in the StD group, which, while consistent with the epidemiology of depression, limits generalizability and prevented adequate examination of sex-specific glymphatic patterns.

### 4.7 Conclusion

This study provides pioneering multimodal neuroimaging evidence of glymphatic system dysfunction in adolescents with subthreshold depression. Our findings reveal a distinct pattern of lateralized glymphatic impairment, strong associations with transdiagnostic features like sleep disturbance, and the successful application of an integrated Mahalanobis distance metric to capture the multifaceted nature of this dysfunction. These results offer crucial neurobiological support for a dimensional view of depression, demonstrating that significant pathophysiology is present even before the criteria for a major depressive disorder are met. Our study directly addresses this gap by demonstrating measurable biological dysfunction in individuals who do not meet full MDD criteria yet experience significant clinical burden.

## Supplementary material

**Table S1.**
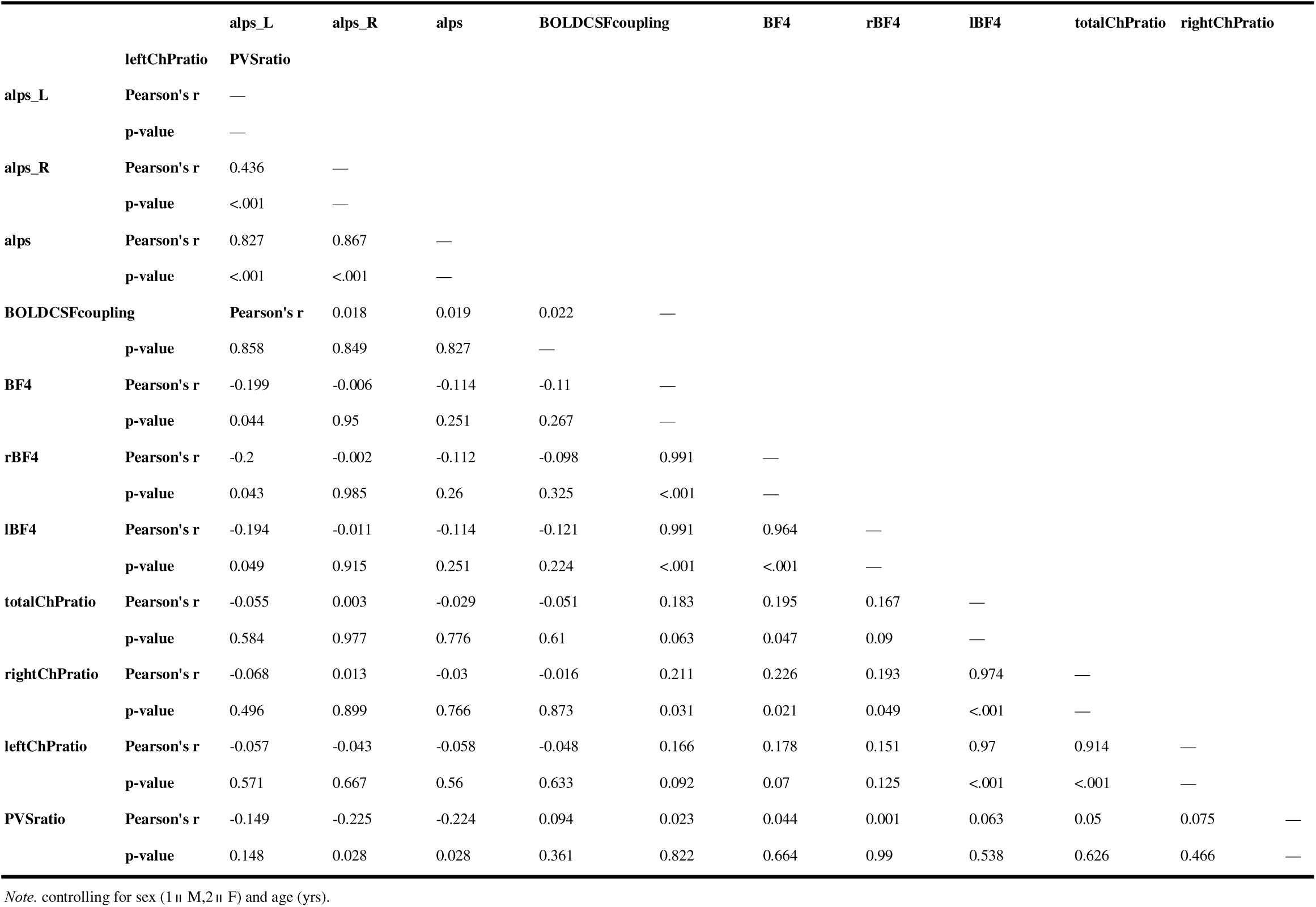
Partial Correlation among glymphatic indices controlling for Age and Gender. Note: alps_L = left hemisphere aperiodic slope; alps_R = right hemisphere aperiodic slope; alps = bilateral aperiodic slope; hurst = Hurst exponent; BOLDCSFcoupling = BOLD-CSF coupling; BF4 = bilateral basal forebrain cholinergic integrity; BF4_R = right basal forebrain cholinergic integrity; BF4_L = left basal forebrain cholinergic integrity; totalChPratio = total choroid plexus ratio; rightChPratio = right choroid plexus ratio; leftChPratio = left choroid plexus ratio; PVSratio = perivascular space ratio. *p < 0.05, **p < 0.01, ***p < 0.001.

**Table S2.** Results of Analysis of Covariance (ANCOVA) for Glymphatic Biomarkers.

| Variable | Gender Effect |  | Age Effect |  | Group Effect |  |
| --- | --- | --- | --- | --- | --- | --- |
|  | F | p | F | p | F | p |
| BOLD-CSF coupling | 0.59 | .445 | 1.83 | .179 | 4.14 | .044* (HC < StD) |
| BF4 | 0.84 | .363 | <0.01 | .982 | 3.17 | .078 |
| BF4_R | 1.17 | .282 | 0.01 | .907 | 3.39 | .069 |
| BF4_L | 0.54 | .466 | <0.01 | .943 | 2.85 | .094 |
| Total ChP ratio | 1.43 | .234 | <0.01 | .992 | 1.93 | .168 |
| PVS ratio | 0.26 | .614 | 0.19 | .668 | 0.07 | .788 |
| Hurst exponent | 3.00 | .087 | 0.17 | .684 | 1.97 | .164 |
| ALPS_L | 0.16 | .687 | 2.34 | .129 | 9.12 | .003** (HC > StD) |
| ALPS_R | 0.25 | .617 | 0.03 | .871 | 0.24 | .623 |
| ALPS | 0.24 | .624 | 0.86 | .355 | 3.77 | .055 |

*Note.* Group: 1 = StD group, 2 = Healthy control group; Gender: 1 = Male, 2 = Female.
BOLD-CSF = Blood oxygen level-dependent cerebrospinal fluid; BF4 = Fourth ventricle;
BF4\_R = right fourth ventricle; BF4\_L = left fourth ventricle; ChP = choroid plexus; PVS =
perivascular space; ALPS = analysis along the perivascular space; StD = Subthreshold
Depression; HC = Healthy controls.
\* $p < .05$ . \*\* $p < .01$ . Direction of significant group differences shown in parentheses.

**Table S3.**
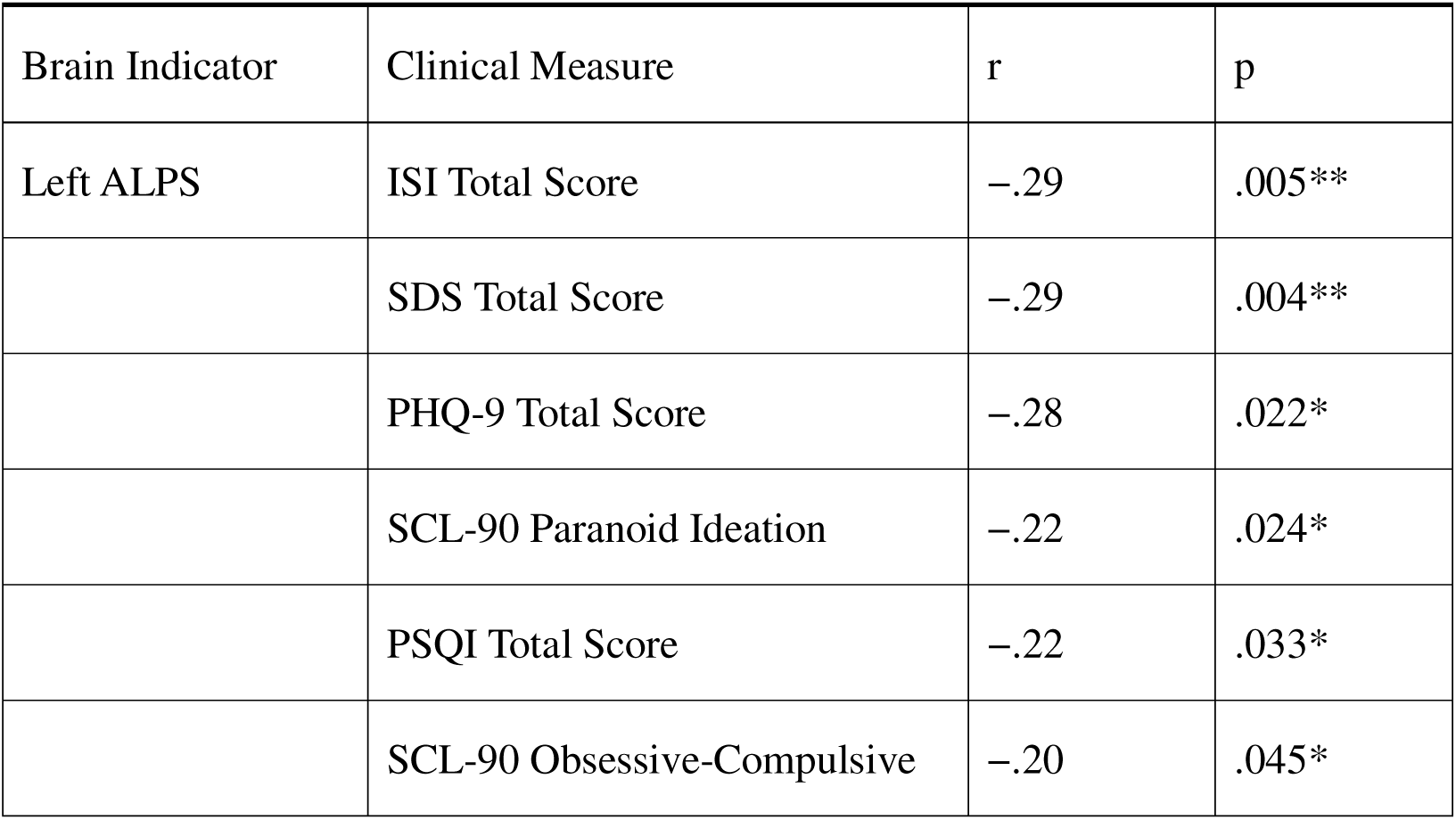
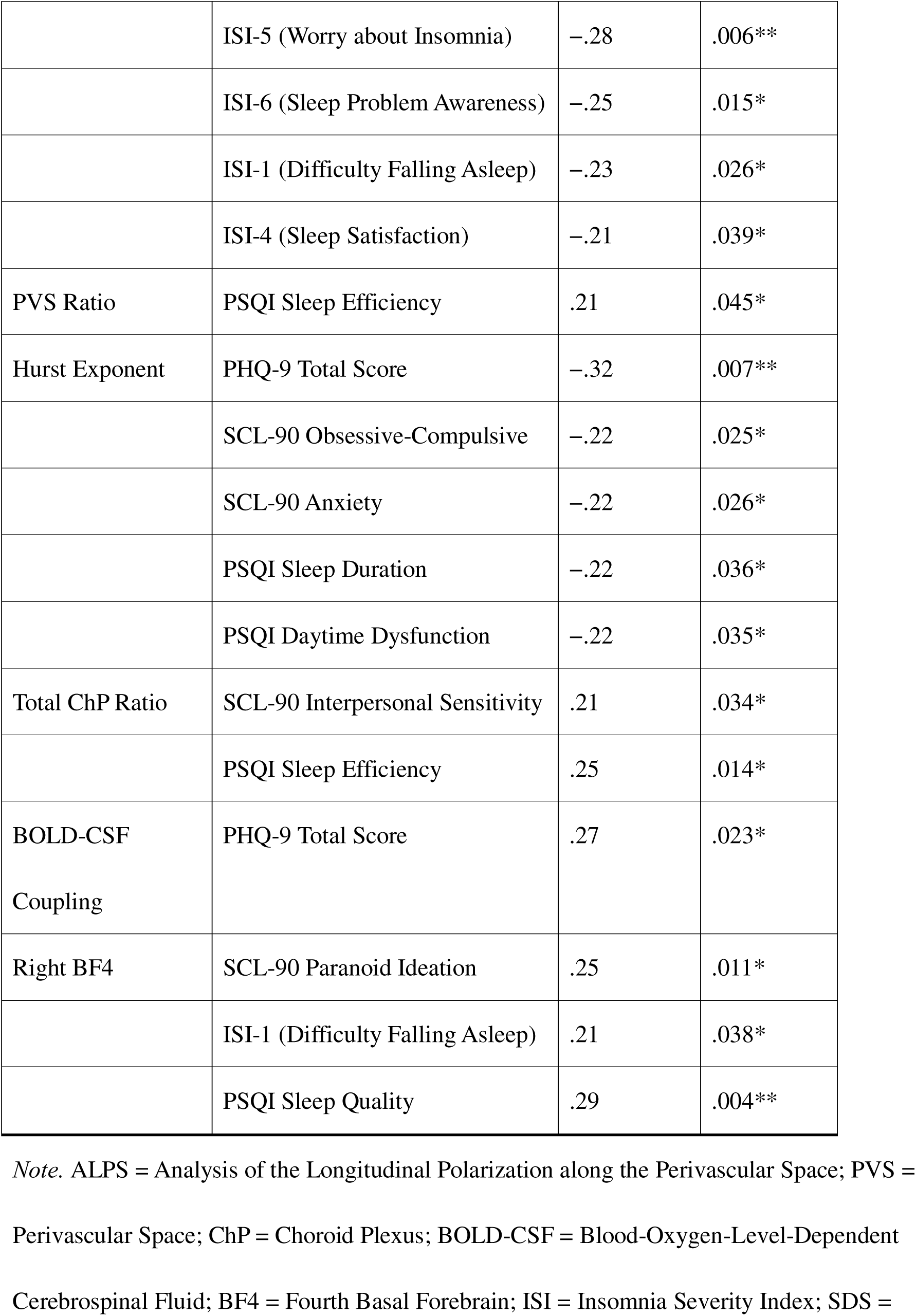
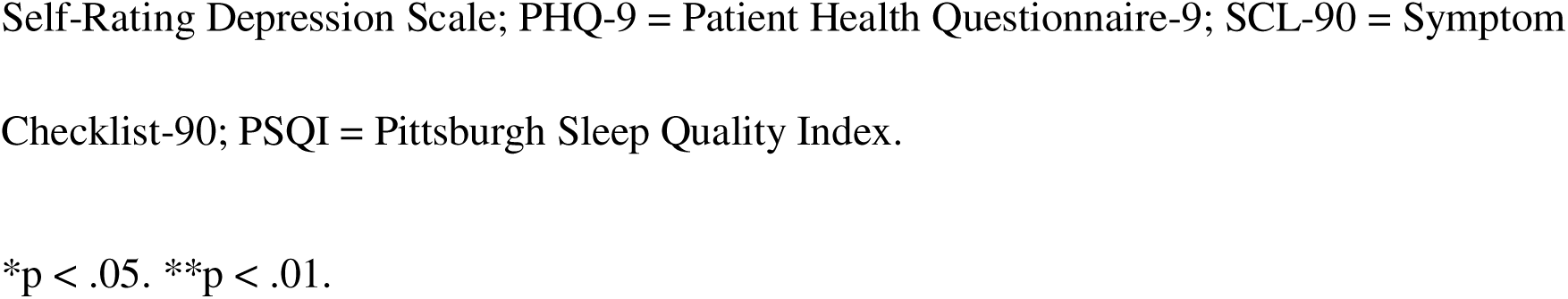
Partial Correlations Between Brain Indicators and Clinical Measures.

## Funding Source

This work was supported by: Special Research Program of Traditional Chinese Medicine Science and Technology, Guangdong Provincial Hospital of Chinese Medicine (Grant No. YN2023MS15), Zhuhai Science and Technology Program for Social Development (Grant No. 2420004000315), Traditional Chinese Medicine Research Project of Guangdong Provincial Administration of Traditional Chinese Medicine (Grant No. 20251351); Programs of University of Macau (MYRG2022-00054-FHS, MYRG-GRG2023-00038-FHS-UMDF, and MYRG-GRG2024-00259-FHS), and the Macao Science and Technology Development Fund (FDCT 0014/2024/RIB1).

## Statement of interest conflict

The authors declare no competing financial interests or personal relationships that could have appeared to influence the work reported in this paper.

## Data availability

De-identified data supporting the findings of this study are available from the corresponding authors upon reasonable request. Due to ethical and legal restrictions related to participant privacy and informed consent provisions, the raw neuroimaging data cannot be deposited in a public repository. However, we are committed to sharing data with qualified researchers for the purpose of replicating or extending our findings, subject to:

1. Approval by the institutional review board of the University of Macau and Guangdong Provincial Hospital of Chinese Medicine
2. Execution of a formal data sharing agreement
3. Confirmation that the proposed use complies with participant consent and local data protection regulations

## Author contributions

Conceptualization: R.W., Z.L.

Methodology: R.W., Q.G.

Software: Q.G., R.W.

Validation: Q.G., R.W.,U.S., Z.W.

Formal Analysis: R.W., Q.G., J.Z.

Investigation: Z.L., Y.Z.

Resources: Z.Y., J.C.

Data Curation: Z.L., C.Z.

Writing – Original Draft: Q.G.

Writing – Review & Editing: Z.Y., J.C.

Visualization: R.W.

Supervision: A.M., Z.Y., J.C.

Project Administration: L.M., Z.Y., J.C.

Funding Acquisition: Z.Y., J.C.

## Notes

### Competing Interest Statement

The authors have declared no competing interest.

### Author Declarations

The Ethics Committee of Zhuhai Hospital of Guangdong Provincial Hospital of Chinese Medicine

